# Comparative Analysis of Adverse Event Reporting on Reddit and the FDA Adverse Event Reporting System (FAERS)

**DOI:** 10.1101/2024.08.13.24311929

**Authors:** Catherine C. Shoults, Dana Abulez, Matthew Nowak, Alasiah Bledsoe, Nathan Dormer, Maryam Y. Garza, Jonathan Bona, Thomas Powell, Corey J. Hayes

**Affiliations:** Department of Biomedical Informatics, College of Medicine, University of Arkansas for Medical Sciences, Little Rock, Arkansas, USA; College of Medicine, University of Arkansas for Medical Sciences, Little Rock, Arkansas, USA; Information Systems and Operations Management Concentration, Goizueta Business School, Emory University, Atlanta, Georgia, USA; Adare Pharma Solution, Lenexa, Kansas, USA; Institute for Digital Health and Innovation, College of Medicine, University of Arkansas for Medical Sciences, Little Rock, Arkansas, USA; Central Arkansas Veterans Healthcare System, Eugene J. Towbin Healthcare Center, North Little Rock, Arkansas, USA

## Abstract

This study explores the potential of Reddit as a pharmacovigilance data source by comparing its adverse event reports related to mental health drugs with those from the FDA Adverse Event Reporting System (FAERS). Using data mining techniques, we annotated 1,000 Reddit posts to identify drug-adverse event pairs, which were then compared to FAERS data. Significant differences were noted in the frequency and types of adverse events reported on each platform. For instance, Reddit showed higher reports of sexual dysfunction and cognitive disorders for certain drugs, which were less frequently reported in FAERS. This disparity suggests that Reddit could capture a different demographic or more candid discussions, potentially influenced by its pseudonymous nature. The findings indicate that social media platforms like Reddit can provide valuable insights into real-world drug effects, complementing traditional pharmacovigilance systems. This study highlights the need for further research to integrate social media data to enhance drug safety monitoring.

## Introduction

Pharmacovigilance, the study of a drug’s safety and adverse event profile, is a crucial aspect of drug development and patient safety. The Food and Drug Administration (FDA) provides formal reporting systems, such as the FDA Adverse Event Reporting System (FAERS) for drug manufacturers, clinicians, and consumers to report adverse events (1). The consumer reporting system suffers from bias in underreporting (2), who reports (3), how complete the reports are (4), notoriety bias (a type of selection bias in which adverse events more well known are more likely to be reported) (5), and what adverse events are considered worthy of reporting (6). Social media data represent an exciting pharmacovigilance data source because of the ability to uncover adverse events sooner (7), receive reports from a novel population (8), and understand real-world use of drugs (9). While many studies have examined the methodology for extracting pharmacovigilance posts using machine learning and natural language processing (NLP) (10–16), few studies have provided characterization and quantification of what adverse events are found in social media. Of the research that characterizes the medical effects social media users ascribe to their prescription drugs use, the most popular source of data has been the website, Twitter^1^ (17,18). However, the free bulk data available from Twitter’s Academic application programming interface (API) has been suspended (19) and novel sources of social media pharmacovigilance data are needed. Reddit is an ideal source of pharmacovigilance data, especially around drugs that deal with mental health, because of its pseudonymous environment and large character base allowing free-flowing dialogue (20,21). Boettcher’s review of the literature on social media use in healthcare research found 54 studies using the Reddit platform discussed mental health. Of those, 94 percent were about depression, and 43 percent researched anxiety (21). Despite its rich text environment and discussion of mental health, Reddit is underused as a pharmacovigilance data source perhaps because of its diverse topic areas, long character base, and lack of peer reviewed literature (22).

The objective of this study was two-fold: (1) to evaluate the feasibility of mining Reddit data on mental health drug use and adverse events and (2) to compare Reddit-derived data to FAERS- derived data to evaluate whether Reddit-derived data capture the essential elements needed for FAERS reporting. This paper expands the literature by recording the adverse events found in Reddit and quantifying how this social media pharmacovigilance source compares to the adverse events voluntarily reported by consumers directly to the FDA via FAERS.

## Methods

### Reddit Data

The Reddit API created by Jason Baumgartner, Pushshift.io, was used to create a Reddit corpus (23). The top 15 psychoactive drugs prescribed in the United States (24) were used as search terms due to their higher rates of adverse events and more common use by younger populations. Search terms including both the branded name and the active pharmaceutical ingredient (see Table 1 for each drug’s rank, brand name, active ingredient, and indication). This ranking was provided by IQVIA (formerly IMS Health and Quintiles), a leading global provider of advanced analytics and contract research services to the healthcare and clinical trial industry. IQVIA prescription data covers 93 percent of outpatient prescriptions.^2^ In addition to the top 15 drug names and ingredients, misspellings were included as Pushshift.io search terms. Misspellings were derived using word2vec.

**Table 1:** List of top 15 psychoactive drugs as identified by IQVIA. Rank is based on number of prescriptions in the United States. Indication is from the FDA label (package insert) of the drug.

| **IQVIA Rank** | **Brand Name** | **Active Ingredient** | **Indication** |
| --- | --- | --- | --- |
| **1** | Zoloft | sertraline | depression |
| **2** | Xanax | alprazolam | anxiety |
| **3** | Lexapro | escitalopram | depression |
| **4** | Desyrel | trazodone | anxiety, depression |
| **5** | Wellbutrin | bupropion | depression |
| **6** | Adderall | dextroamphetamine,<br>amphetamine | ADHD |
| **7** | Prozac | fluoxetine | depression |
| **8** | Celexa | citalopram | depression |
| **9** | Cymbalta | duloxetine | depression |
| **10** | Ativan | lorazepam | anxiety |
| **11** | Effexor | venlafaxine | depression |
| **12** | Seroquel | quetiapine | bipolar disorder, depression |
| **13** | Lamictal | lamotrigine | bipolar disorder |
| **14** | Concerta | methylphenidate | ADHD |
| **15** | Kapvay | clonidine | ADHD |

One hundred thousand (100,000) random Reddit posts were pulled from the full Reddit corpus. This sample of posts were fed into Prodi.gy Bulk, which takes the textual posts and generates their embeddings using the sentence transformer model paraphrase-MiniLM-L6-v2. Paraphrase-MiniLM-L6-v2 is a pre-trained language model developed by Hugging Face (25).

The embeddings were transformed from multi-dimensional embeddings to two-dimensional space using Uniform Manifold Approximation and Project (UMAP). This 2D representation allows for an interactive visualization to be created, where each dot represents a Reddit post, and hovering over the dot allows the user to see the post’s text. The dots were color coded based on if they contained the phrase “adverse event” or “side effect”, and clusters with these terms were manually reviewed to determine if the cluster of posts contained discussion of pharmacovigilance posts.

Clusters of interest were downloaded from the Prodi.gy Bulk interface and annotated as discussing pharmacovigilance. Posts were labeled to conform to the definition of a safety report outlined in the *Guideline on Clinical Safety Data Management: Data Elements for the Transmission of Individual Case Safety Reports*: a safety report is defined to require an identifiable patient, an identifiable reporter, a suspect drug, and an adverse event (26,27). In order to ensure that all safety reports met this definition, only posts where the poster was writing about their own personal experience were included. This approach has been used previously to best conform to the International Council for Harmonization (11) guidelines.

Three annotators independently extracted 1000 drug-adverse event pairs. Posts were studied to find relationships between the Redditor’s drug use and listed adverse event. Annotators were instructed to look for causal language (e.g., “because”, “caused”, “led to”, “made me”) linking a drug to an adverse event. Table 2 shows example text linking an adverse event to a listed drug. “Drug” was defined as any drug name, including branded name, generic name, and misspellings (drug classes were not included). “Adverse event” was defined as any attributed event described as due to the drug. If an adverse event could be attributed to more than one drug, each drug-adverse event pair was listed individually (polypharmacy is outside the scope of this research). Indication or disease states were recorded when the author stated they were taking the drug for a stated indication, listed their current disease state, or when the author posted to a disease-specific subreddit.

**Table 2:** Synthetic Reddit posts showing how annotations were recorded. Note that Reddit Post with ID 2 is duplicated because two adverse events were discussed in the post.

| ID | Reddit Post | Drug | Adverse Event |
| --- | --- | --- | --- |
| 1 | I was on Zoloft but quit because I couldn't sleep. | Zoloft | couldn't sleep |
| 2 | I've been on Wellbutrin and Lamictal. I really loved Lamictal but it stopped working and I got a rash. | Lamictal | stopped working |
| 2 | I've been on Wellbutrin and Lamictal. I really loved Lamictal but it stopped working and I got a rash. | Lamictal | Rash |
| 3 | Yeah, Lexapro definitely gave me brain zaps. I've heard other people get tremors but that didn't happen to me. | Lexapro | Brain zaps |

If multiple drugs were mentioned in combination with an adverse event without causal language to just one drug, the drug-to-adverse event pair was listed for each drug. If the same adverse event was listed multiple times in a post, it was only recorded once. If the drug was listed as ineffective, this was documented as an adverse event. Adjudication was performed between the annotators to reconcile differences.

### FAERS Data

FAERS was accessed via the FDA’s public dashboard (28). The top five most common drugs from the Reddit corpus were used to query the FAERS dashboard. The query was limited to consumer reporters. After obtaining all adverse event reports from FAERS, the count of the five most reported adverse events and the total number of case reports per drug were recorded for comparison to the Reddit results.

### Normalization

Drugs were normalized to their branded name. A dictionary of brand name, generic, and common misspellings was used to standardize the drugs to their branded name. Adverse events were normalized to the FAERS Side Effect Resource (SIDER) Preferred Term. A medical doctor reviewed these mappings for clinical plausibility (see Appendix 1 to view mapping of adverse event to FAERS term and clinical notes).

### Statistical Analysis

Python 3.8.5 was used to analyze these data. Pandas was used to calculate relative frequencies and examine adverse event descriptive statistics (29). The Reddit data and FAERS data were merged using an inner join on the drug and FAERS-normalized adverse event terms. The SciPy Stats package was used to compare the FAERS adverse events to the Reddit adverse events using Chi-Square tests (30). The null hypothesis was that there was no statistical significance between FAERS adverse events and Reddit adverse events. This hypothesis was tested using a p-value set to 0.05. Visualizations were created with Matplotlib (31) and Seaborn (32).

## Results

The Reddit data included 1000 drug-adverse event pairs spanning from 2010 to 2022 (see Table 3 for the distribution by year). The most common drug in the Reddit posts was Zoloft followed by Lexapro, Effexor, and Lamictal (see Figure 1). Zoloft had 231 drug mentions, Lexapro had 229 drug mentions, Effexor had 178 drug mentions, Lamictal had 148, and Wellbutrin had 114 drug mentions.

**Figure 1:**
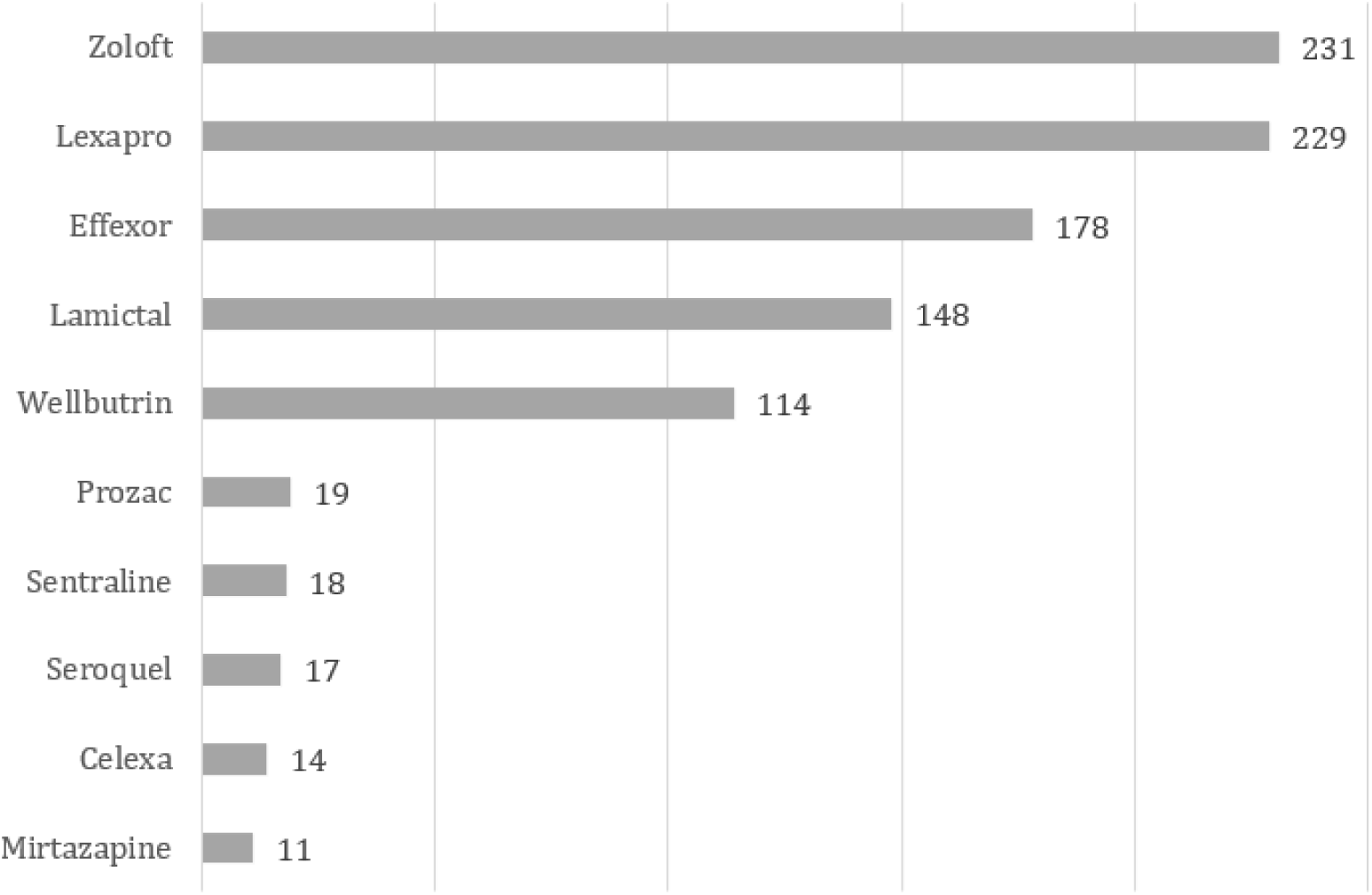
Most common drug names found in the Reddit posts along with the number of mentions of the drug name. Drug names represent mentions of drugs in their branded form, generic name, and misspellings. Note: “Sentraline” may be a misspelling of “Sertraline” meaning these could be under the heading of “Zoloft”. The two were kept separate in this analysis.

**Table 3:** Number of Reddit posts by the year that the post was made on the platform.

| **Post Year** | **Post Count** |
| --- | --- |
| 2022 | 225 |
| 2021 | 271 |
| 2020 | 191 |
| 2019 | 120 |
| 2018 | 37 |
| 2017 | 49 |
| 2016 | 45 |
| 2015 | 32 |
| 2014 | 6 |
| 2013 | 5 |
| 2012 | 3 |
| 2011 | 10 |
| 2010 | 6 |

Reddit subreddits by drug type are noted in Figure 2. The subreddits reflect the disease states that the Redditors discussed in their posts. The top subreddits were often named for the drug’s branded or generic name or common diseases associated with the drug. The top subreddit for Effexor was r/effexor. Lamictal’s top subreddit was r/bipolar, and Lexapro’s top subreddit was r/lexapro. Wellbutrin was most commonly found in r/bupropion and Zoloft’s in r/zoloft. The disease states associated with each drug were often depression, anxiety, and bipolar disease. Depression was the most common disease mentioned for Effexor and Wellbutrin drug-adverse event pairs. Anxiety was the top disease mentioned for Lexapro and Zoloft. Bipolar disease was the top disease listed for Lamictal.

**Figure 2:**
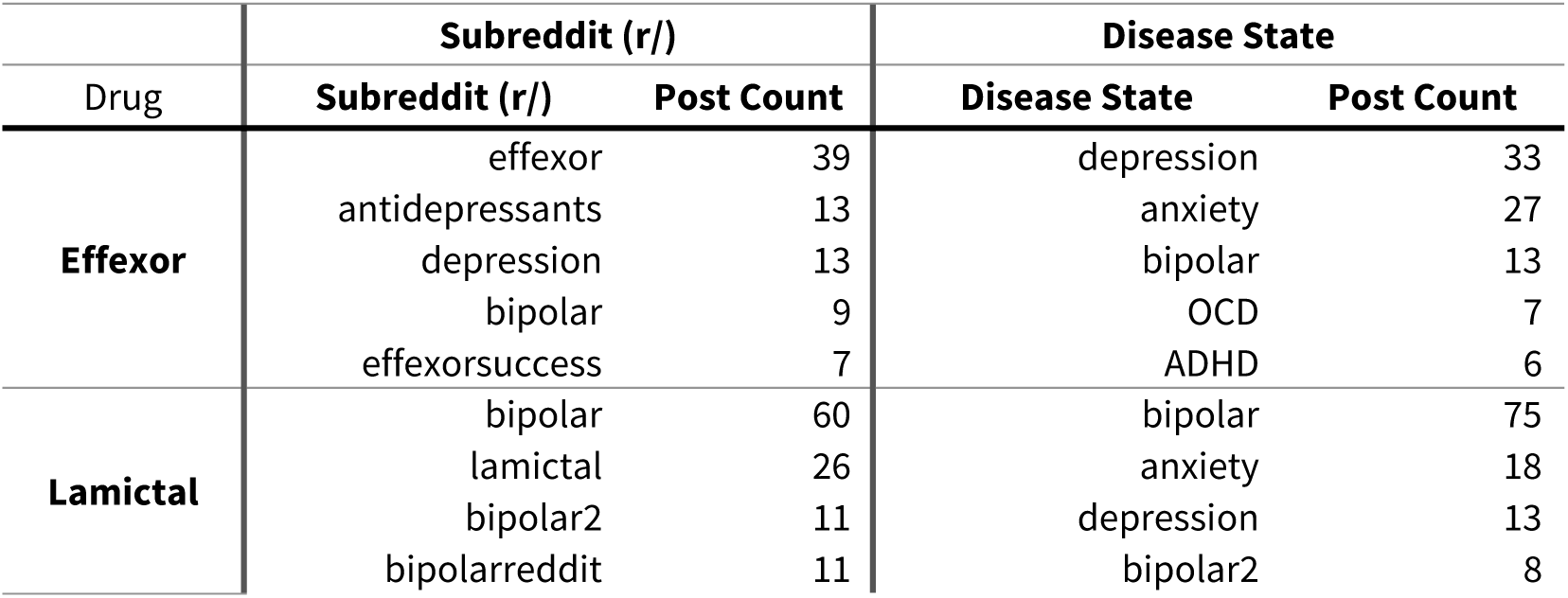
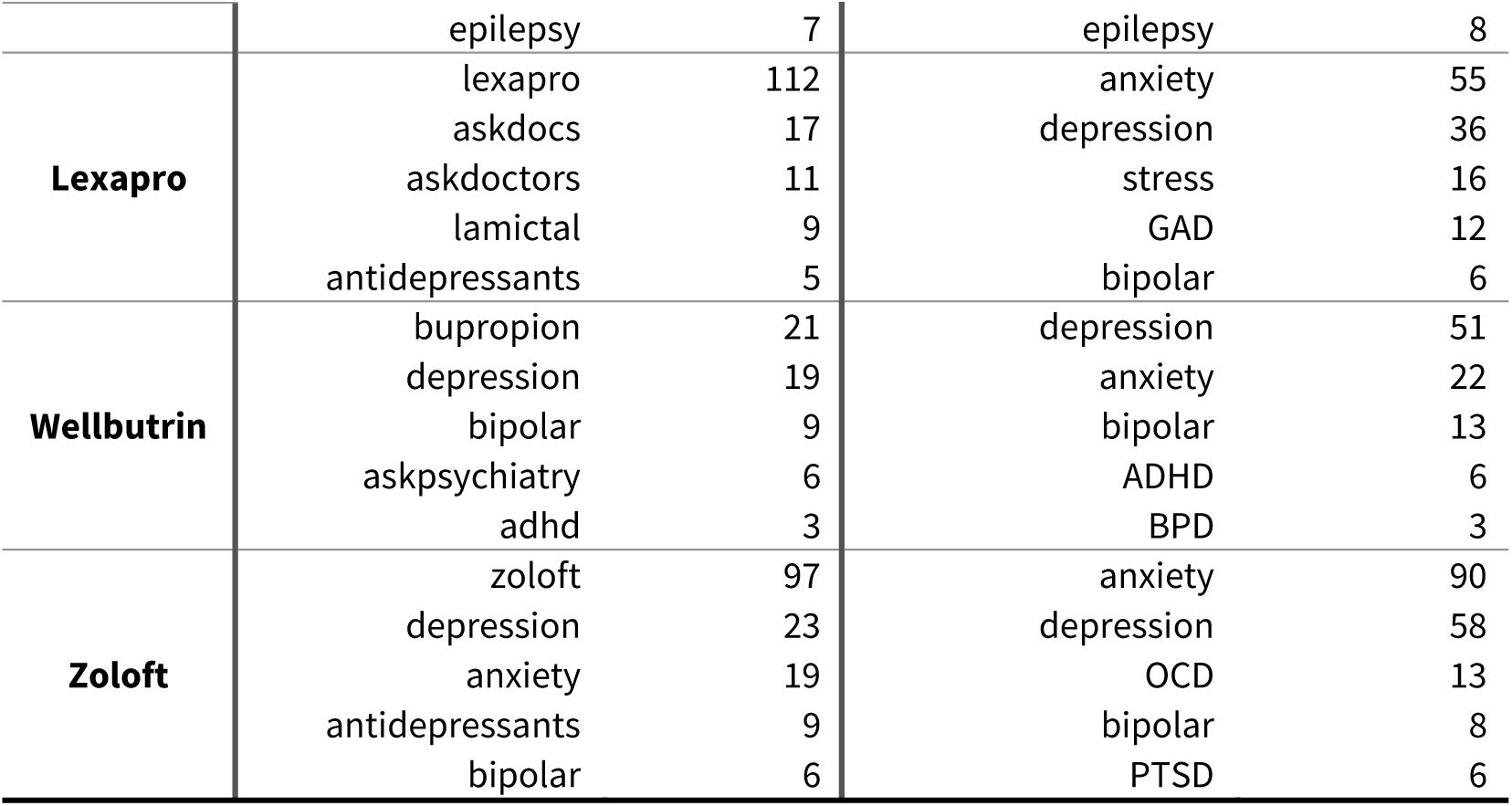
Count of subreddits and disease states associated with drugs that were mentioned in drug-adverse event pairs.

### FAERS Data Results

A total of 62,787 reports were obtained from the FAERS dashboard, spanning from 2010 through 2023. The top five most common adverse events for each drug are listed in Table 4. The most common drug found in FAERS was Zoloft, with 23,732 adverse event reports. This was followed by Lamictal, with 16,043 reports, and Wellbutrin, with 11,406. Effexor had 7,576 reports, and Lexapro had 4,030 reports. Zoloft’s most commonly reported adverse events were the Drug was Ineffective, Insomnia, and Anxiety. Lamictal’s most common adverse event was Rash, Seizure, and Drug Ineffective. Wellbutrin had Drug Ineffective as its most common adverse event, followed by Insomnia and Anxiety. Effexor’s top adverse event was Drug Withdrawal Syndrome, Dizziness, and Drug Ineffective. Finally, Lexapro’s most common adverse events were Drug Ineffective, Anxiety, and Depression.

**Table 4:** Five most common drugs in the Reddit gold standard posts and their corresponding FAERS drug counts and top five adverse events.

| **Effexor**<br>**(7,576)** | **Lamictal**<br>**(16,043)** | **Lexapro**<br>**(4,030)** | **Wellbutrin**<br>**(11,406)** | **Zoloft**<br>**(23,732)** |
| --- | --- | --- | --- | --- |
| Drug Withdrawal Syndrome (902) | Rash (2,575) | Drug Ineffective (474) | Drug Ineffective (1,272) | Drug Ineffective (1,919) |
| Nausea (683) | Seizure (1,571) | Anxiety (395) | Seizure (828) | Insomnia (1,567) |
| Dizziness (667) | Drug Ineffective (1,322) | Depression (383) | Insomnia (800) | Anxiety (1,553) |
| Drug Ineffective (555) | Headache (825) | Suicidal Ideation (345) | Headache (648) | Dizziness (1,402) |
| Headache (548) | Pruritus (798) | Nausea (314) | Depression (621) | Nausea (1,376) |

### Reddit Data Results

The most common drugs mentioned in the 1000 Reddit posts were Zoloft, Lexapro, Effexor, Lamictal, and Wellbutrin, as can be seen in Table 5. The most commonly mentioned of Zoloft’s 231 adverse events were Fatigue, Apathy, and Sexual Dysfunction. Of Lexapro’s 229 adverse events, Sexual Dysfunction, Apathy, and Cognitive Disorder were the most common. Of Effexor’s 178 adverse events, Sexual Dysfunction, Fatigue, and Paranesthesia were the most common. Of Lamictal’s 148 adverse events, Cognitive Disorder, Anxiety, and Fatigue were the most common. Finally, of Wellbutrin’s 114 adverse events, Anxiety, Fatigue, and Mania were the most common.

**Table 5:** Five most common drugs in the Reddit gold standard posts and their corresponding drug counts and top five adverse events.

| **Effexor**<br>**(n=178)** | **Lamictal**<br>**(n=148)** | **Lexapro**<br>**(n=229)** | **Wellbutrin (n=114)** | **Zoloft**<br>**(n=231)** |
| --- | --- | --- | --- | --- |
| Sexual Dysfunction<br>(20) | Cognitive Disorder<br>(11) | Sexual Dysfunction<br>(21) | Anxiety (8) | Fatigue (22) |
| Fatigue (10) | Anxiety (9) | Apathy (18) | Fatigue (6) | Apathy (18) |
| Paranesthesia (6) | Fatigue (8) | Cognitive Disorder<br>(11) | Mania (6) | Sexual Dysfunction<br>(15) |
| Weight Increased (6) | Apathy (5) | Anxiety (10) | Cognitive Disorder<br>(5) | Anxiety (9) |
| Anxiety (4) | Irritability (5) | Fatigue (8) | Irritability (5) | Insomnia (9) |

### Drug and Adverse Event Comparison

The top five drugs in this study, Effexor, Lamictal, Lexapro, Wellbutrin, and Zoloft, were compared across Reddit and FAERS via Chi-Square analysis (Table 6). Reddit’s Effexor reporting rates of Fatigue, Paranesthesia, and Weight Increased were found to be similar between Reddit and FAERS (p-values of 0.054, 0.625, and 0.876, respectively). However, Sexual Dysfunction showed a significantly higher reporting rate on Reddit compared to FAERS, with a p-value of less than 0.001.

**Table 6:** Drug and Adverse Event pairs for top Reddit drugs compared to FAERS using chi-square analysis; FAERS: FDA Adverse Event Reporting System

| Drug Type and FAERS Adverse Event Category | Reddit Frequency Count and Percent | Reddit Sample Size | FAERS Frequency Count and Percent | FAERS Sample Size | Chi-Square | P-value |
| --- | --- | --- | --- | --- | --- | --- |
| **Effexor** |  |  |  |  |  |  |
| Fatigue | 10 (5.6%) | 178 | 233 (3.1%) | 7576 | 3.7 | 0.054 |
| Paranesthesia | 6 (3.4%) | 178 | 311 (4.1%) | 7576 | 0.24 | 0.625 |
| Sexual Dysfunction | 20 (11.2%) | 178 | 54 (0.7%) | 7576 | 203.74 | <0.001 |
| Weight Increased | 6 (3.4%) | 178 | 272 (3.6%) | 7576 | 0.02 | 0.876 |
| **Lamictal** |  |  |  |  |  |  |
| Anxiety | 9 (6.1%) | 148 | 467 (2.9%) | 16043 | 5.16 | 0.023 |
| Apathy | 5 (3.4%) | 148 | 34 (0.2%) | 16043 | 61.19 | <0.001 |
| Cognitive Disorder | 11 (7.4%) | 148 | 98 (0.6%) | 16043 | 102.05 | <0.001 |
| Fatigue | 8 (5.4%) | 148 | 621 (3.9%) | 16043 | 0.92 | 0.336 |
| Irritability | 5 (3.4%) | 148 | 216 (1.4%) | 16043 | 4.5 | 0.034 |
| **Lexapro** |  |  |  |  |  |  |
| Abnormal Dreams | 5 (2.2%) | 229 | 43 (1.1%) | 4030 | 2.42 | 0.120 |
| Anxiety | 10 (4.4%) | 229 | 395 (9.8%) | 4030 | 7.44 | 0.006 |
| Apathy | 18 (7.9%) | 229 | 35 (0.9%) | 4030 | 86.19 | <0.001 |
| Cognitive Disorder | 11 (4.8%) | 229 | 23 (0.6%) | 4030 | 49.02 | <0.001 |
| Depression | 6 (2.6%) | 229 | 383 (9.5%) | 4030 | 12.37 | <0.001 |
| Fatigue | 8 (3.5%) | 229 | 292 (7.2%) | 4030 | 4.66 | 0.031 |
| Insomnia | 7 (3.1%) | 229 | 255 (6.3%) | 4030 | 4.02 | 0.045 |
| Irritability | 6 (2.6%) | 229 | 99 (2.5%) | 4030 | 0.02 | 0.877 |
| Mania | 5 (2.2%) | 229 | 72 (1.8%) | 4030 | 0.19 | 0.661 |
| Sexual Dysfunction | 21 (9.2%) | 229 | 51 (1.3%) | 4030 | 81.47 | <0.001 |
| **Wellbutrin** |  |  |  |  |  |  |
| Anxiety | 8 (7.0%) | 114 | 613 (5.4%) | 11406 | 0.6 | 0.440 |
| Cognitive Disorder | 5 (4.4%) | 114 | 30 (0.3%) | 11406 | 63.34 | <0.001 |
| Fatigue | 6 (5.3%) | 114 | 361 (3.2%) | 11406 | 1.61 | 0.204 |
| Irritability | 5 (4.4%) | 114 | 183 (1.6%) | 11406 | 5.44 | 0.020 |
| Mania | 6 (5.3%) | 114 | 99 (0.9%) | 11406 | 24.14 | <0.001 |
| **Zoloft** |  |  |  |  |  |  |
| Anxiety | 9 (3.9%) | 231 | 1,553 (6.5%) | 23732 | 2.63 | 0.105 |
| Apathy | 18 (7.8%) | 231 | 388 (1.6%) | 23732 | 52.07 | <0.001 |
| Depression | 6 (2.6%) | 231 | 1,367 (5.8%) | 23732 | 4.24 | 0.040 |
| Dizziness | 5 (2.2%) | 231 | 1,402 (5.9%) | 23732 | 5.8 | 0.016 |
| Fatigue | 22 (9.5%) | 231 | 769 (3.2%) | 23732 | 28.3 | <0.001 |
| Insomnia | 9 (3.9%) | 231 | 1,567 (6.6%) | 23732 | 2.73 | 0.099 |
| Mania | 5 (2.2%) | 231 | 249 (1.1%) | 23732 | 2.71 | 0.100 |
| Nausea | 6 (2.6%) | 231 | 1,376 (5.8%) | 23732 | 4.31 | 0.038 |
| Sexual Dysfunction | 15 (6.5%) | 231 | 169 (0.7%) | 23732 | 100.36 | <0.001 |
| Tremor | 5 (2.2%) | 231 | 906 (3.8%) | 23732 | 1.71 | 0.191 |

Moving to Lamictal, several adverse events, including Anxiety, Apathy, Cognitive Disorder, and Irritability, exhibited significant differences between the two datasets, with Reddit reporting higher frequencies for each (p-values of 0.023, <0.001, <0.001, and 0.034, respectively). On the other hand, instances of Fatigue showed no significant difference in reporting rates between the two platforms (p-value=0.336). For Lexapro, Reddit had a higher frequency of reported adverse events for Apathy, Cognitive Disorder, Depression, and Sexual Dysfunction when compared to FAERS (p-values<0.001 for all). However, the rate of Anxiety reporting was significantly lower on Reddit than FAERS (p-value=0.006). Other adverse events, such as Abnormal Dreams, Insomnia, Irritability, and Mania, showed no significant difference between the two platforms (p-values of 0.120, 0.045, 0.877, and 0.661, respectively). In the Wellbutrin Reddit dataset, significant differences were observed in the Reddit dataset compared to FAERS for the reporting rates of Cognitive Disorder, Irritability, and Mania (p-values<0.001, 0.020, and <0.001, respectively). Reporting rates for Anxiety and Fatigue showed no significant differences between the two platforms (p-values of 0.440 and 0.204, respectively).

Lastly, for Zoloft, Redditors reported higher rates of Apathy, Fatigue, and Sexual Dysfunction (p-values<0.001 for all). Additionally, the Reddit sample indicated significant differences in the frequencies of Depression, Dizziness, and Nausea (p-values of 0.040, 0.016, and 0.038, respectively). However, reporting rates for Anxiety, Insomnia, Mania, and Tremor were not significantly different between Reddit and FAERS (p-values of 0.105, 0.099, 0.100, and 0.191, respectively). Of note, Drug Withdrawal Syndrome was among the top five most common adverse events in the FAERS reports for Effexor, Lexapro, and Zoloft, which was not observed in the Reddit reporting rates. For visualization of Reddit and FAERS adverse event rates, please see Appendix 2. These findings highlight the differences in adverse event reporting between Reddit and FAERS and provide valuable insights into drug safety assessment across these platforms. The study results were significant in shedding light on potential variations in drug-related adverse event reporting on different platforms.

## Discussion

Social media provides a view into real-world use of drugs and their associated adverse effects. With the suspension of Twitter’s API, alternative sources should be explored. This study assessed differences between the reporting of adverse events between Reddit and FAERS for the top five most common drugs from the gold standard. Overall, similar types of adverse events for each of the five drugs were found in both reporting platforms (i.e., Reddit and FAERS). However, significant differences were found in some drug-adverse event pairs, as well as differences in the rates of similar adverse events. When examining the top five adverse events for each drug discussed in Reddit, new types of issues top the list. Effexor included the addition of Fatigue, Paranesthesia, Weight Increase, and Anxiety. In addition, Sexual Dysfunction was reported at a much higher rate as compared to FAERS. Reddit’s Lamictal reporting rates showed the addition of Cognitive Disorder, Anxiety, Fatigue, Apathy, and Irritability. Lexapro also included Apathy, Cognitive Disorder, Anxiety, and Fatigue but added in Sexual Dysfunction as the most commonly reported adverse event. Wellbutrin reporting in Reddit had Anxiety, Fatigue, Mania, Cognitive Disorder, and Irritability as the top five most reported adverse events as compared to FAERS. Zoloft exhibits significant dissimilarities between Reddit and FAERS with Fatigue, Apathy, Sexual Dysfunction, Anxiety, and Insomnia topping the adverse event list in Reddit. None of these mentioned adverse events were found in the FAERS top five, underscoring the disparity between the two sources. These results indicate social media platforms like Reddit might provide additional insights into patient experiences with drugs. Rates of adverse event reporting were generally higher among Reddit posts, as compared to FAERS, for most events for the top 5 common drugs. In addition, Reddit contained adverse events not reported to FAERS. This indicates Reddit, and potentially other social media platforms, may be helpful sources to compliment FAERS reporting.

Nguyen’s 2017 study examined Twitter, Reddit, and LiveJournal to understand which social media website’s adverse event rates best reflected the adverse events found in the Side Effect Resource (33). Posts from each platform were filtered to only contain 10 psychotropic drugs of interest, and a lexicon was used to look up adverse events (33). The study found that Reddit was most similar in adverse event rates to SIDER as compared to the other social media platforms (33). A limitation of this work was that a simple mention of an adverse event was used as a proxy for the social media user experiencing and writing about an adverse event caused by a drug. Assuming that mentioning an adverse event in a post directly implies the individual experienced it is a limitation, as such references may merely be speculative or descriptive without confirming an actual occurrence.

Another study, conducted in 2020 by Natter and Michel, examined memantine experiences on Reddit and identified side effects from 136 users (34). This is the only pharmacovigilance research documenting specific adverse events described by Redditors available to date.

Twitter-based research reporting drug and adverse event associations are more common. Zhou and Hultgren’s 2020 study compared Twitter adverse events to the FDA’s adverse events (18). The study used tweets labeled for drug-adverse event pairs for comparison to FAERS and provided detailed analysis of what adverse events were due to hydrocodone/acetaminophen, prednisone, amoxicillin, gabapentin, and metformin (18). Li, et al. also studied and reported on detailed Twitter adverse events as compared to FAERS; however, this study used mentions of a potential adverse event as a proxy for identifying the drug-adverse event pair as causally related (according to the text of the social media post) (35). As noted by the authors, the limitations of this assumption make it difficult to make conclusions from the study (35). O’Connor et al. studied tweets for pharmacovigilance and used 1008 hand-annotated tweets to understand the adverse event profiles of 10 drugs of interest. (36). While the study did not compare to the FDA’s consumer reported adverse events, it did list adverse effects and their associated frequency in the tweets (36). Smith et al. used Machine Learning to extract 2617 potential adalimumab adverse events from Twitter and manually annotated to find 801 true adverse events, which were manually mapped to UMLS concept IDs (17). True adverse events were compared to adverse events documented in FAERS and the researchers found moderate agreement between Twitter and the FDA (17).

This study builds on these methods by applying a similar approach to compare Reddit to FAERS. With the suspension of Twitter’s Academic API, the use of social media pharmacovigilance with Reddit data has become even more vital. The evaluation of psychotropic drugs provides novel insights into how Redditors describe the effects of their mental health drugs. This study expands the current literature both by providing Reddit data and by focusing on the drugs that younger, male users may use--a demographic that is less represented in FAERS--Redditors skew young and male (37). The results show differences in reporting to the FDA, especially in the area of Sexual Dysfunction. This could be due to the relative anonymity of Reddit as compared to direct reporting or talking to a clinician. Reddit has been shown to be a place for discussion about mental health (20) and provides a community to discuss shared experiences (38). As noted by Correia et al. in their review of mining social media, clinicians consistently under report depression and pain and rarely use screening tools for adverse events, providing a strong opportunity for social media to afford insights into the patient experience (39). It is also possible that the younger, male user base cares more about the adverse events discussed in Reddit as compared to the older, female user base of the traditional reporting methods. To gain further clarity on this matter, future research could focus on comprehending the demographics of Redditors who post about pharmacovigilance events. Such investigations would offer valuable insights into the differing perspectives and priorities surrounding adverse event discussions across Reddit and FAERS user cohorts.

## Limitations

This study’s findings need to be interpreted in light of several important limitations. The use of causal language to link drug to adverse event is subjective and could introduce bias or inconsistencies in the annotation process. The percent agreement for the normalized terms was 46 percent; however, this was exact match and many of the discrepancies were due to differences in including adjectives or replicating the same adverse event. The subjective nature of human written speech continues to be an issue when mining social media. In addition, hand normalization to the FDA preferred terms could introduce errors or variation into the recorded adverse event to drug relationships. These mappings were reviewed by a clinician to help mitigate this limitation and the mappings can be found in Appendix 1.

Polypharmacy was considered out-of-scope for this research. Many posts contained Redditors who ingested many types of drugs over time or at the same time. This means that an important part of the clinical profile of the Redditors was lost. Future research should integrate polypharmacy into the scope and learn how the real-world use of multiple medications affects adverse event rates.

Chi-square test does not establish causality and should be interpreted as associations and differences, not direct cause-effect relationships. In addition, the Redditor associating an adverse event with their medications does not infer causality. The Redditors are assumed to have a lack of clinical confirmation regarding the accuracy, severity, and causality of the adverse events.

These data were collected using unsupervised Machine Learning. The technique used 100,000 randomly selected posts from a large Reddit corpus, but the 1,000-post subset that was collected may not represent all mental health pharmacovigilance posts in Reddit. The use of a clustering algorithm on word embeddings may have focused the study on similar posts, thereby skewing to that use of language. This limits the generalizability of these findings.

Finally, social media provides a unique platform to peek into the lives of online users; however, the potential for misinformation is always lurking. Anonymity is a “two-edged sword”, with the potential for more vulnerable, open communication offset by mistruths and exaggeration. Despite these limitations, this study contributes to the validation of Reddit as a pharmacovigilance data source and the potential for adding the voices of Redditors to the understanding of drug safety profiles.

## Conclusion

This study demonstrates the feasibility of using Reddit as a pharmacovigilance tool to supplement data collected from FAERS and is particularly rich in capturing adverse event data on population groups are underrepresented in FAERS (e.g., young males). Therefore, Reddit could be used in the future to better estimate adverse event profiles for medications in real-time, which may be particularly important for newly approved medications.

The Reddit pharmacovigilance data analysis identified significant differences from the consumer reported events. The five psychotropic drugs included in the analysis—Effexor, Lamictal Lexapro, Wellbutrin, and Zoloft—encompassed adverse events in areas such as Sexual Dysfunction and Cognitive Disorder that were reported more commonly than what was found in FAERS. Lamictal showed significantly less reporting of Rash in Reddit, which may be indicative of a clinician’s bias to report known relationships between a drug and adverse event. Similarities between the platforms point to consistent issues with use of these drugs (or at least, consistent reporting of issues). Wellbutrin’s profile of Anxiety and Fatigue showcase the similarities between the pharmacovigilance reporting platforms.

Future research in this field should focus on replicating the research with a larger and more diverse dataset. While caution should be used applying rules-based or Machine Learning approaches to infer association between mention of a drug and mention of an adverse event, large scale annotation of relationships will provide additional insights into the distribution and frequency of adverse events in psychotropic medication use as mentioned on Reddit. Our hope is that this study demonstrates the value of Reddit as a complementary data source for pharmacovigilance. Further developing the tools and data from Reddit social media pharmacovigilance can enhance our understanding of drug safety and improve patient care.

## Data Availability

All data produced in the present study are available upon reasonable request to the authors.

## Acknowledgments

This work was generously supported by the National Science Foundation, Award No. OIA- 1946391 Data Analytics that are Robust and Trusted (DART): From Smart Curation to Socially Aware Decision Making.

## Appendix 1 Normalization to FAERS Terms

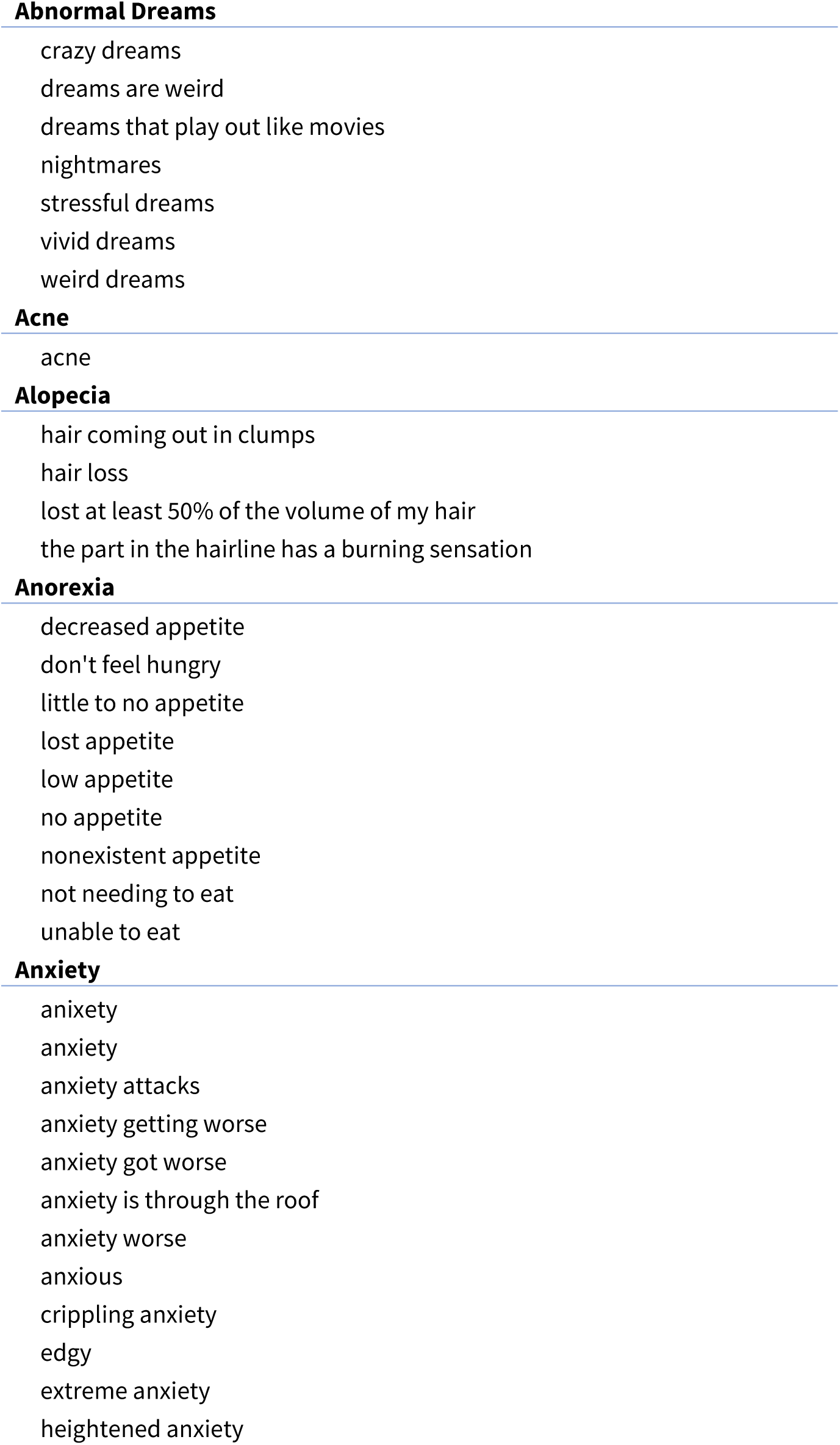
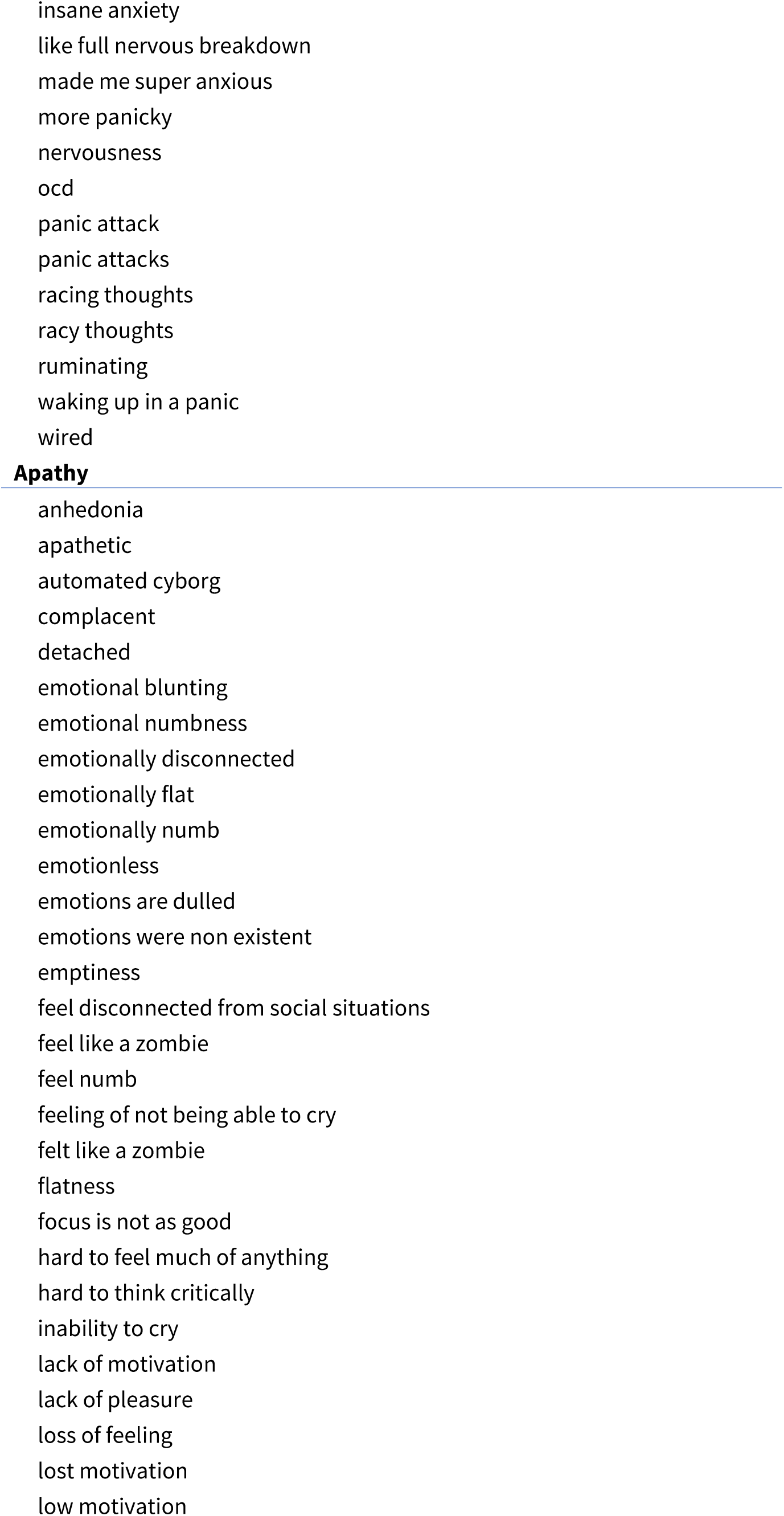
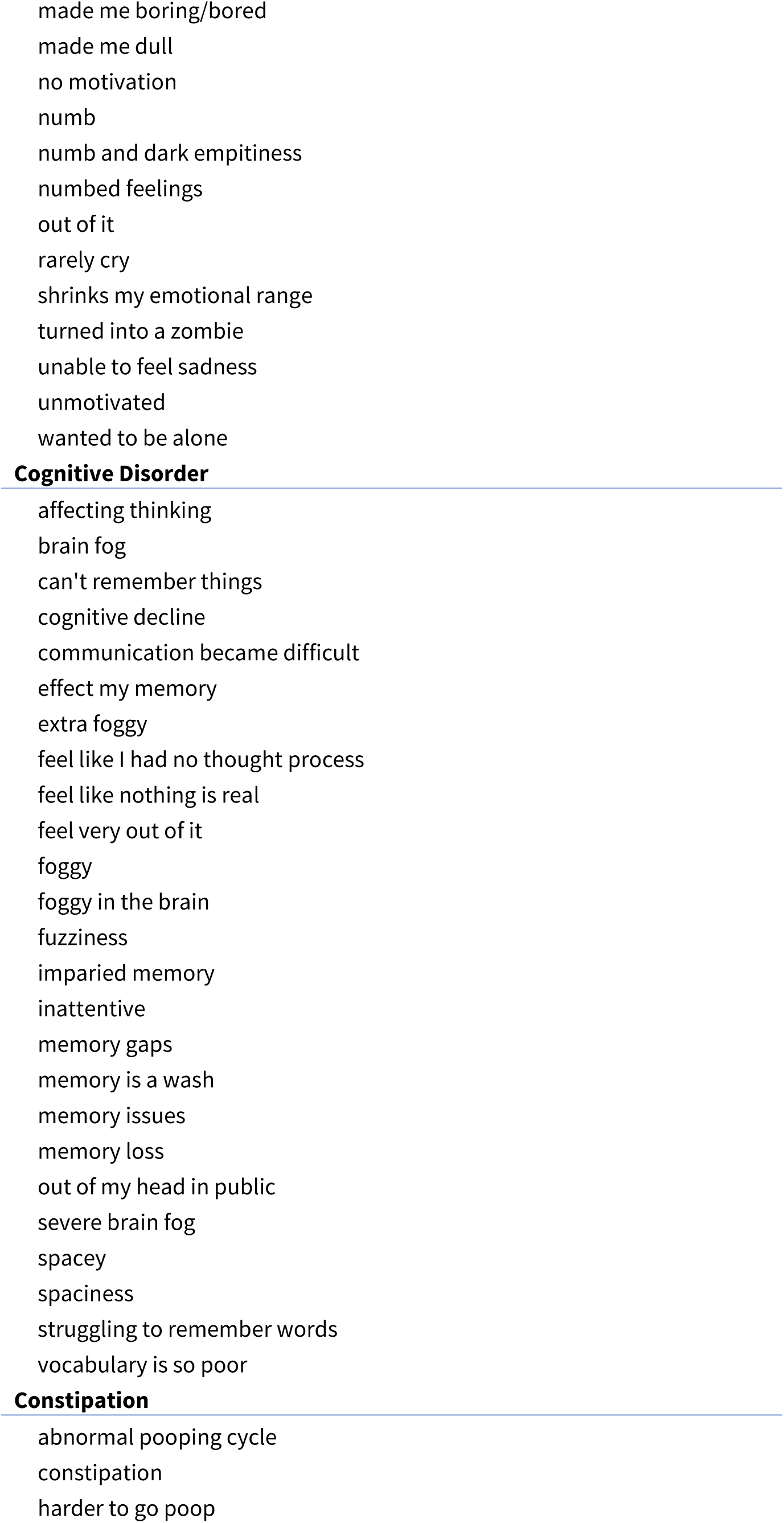
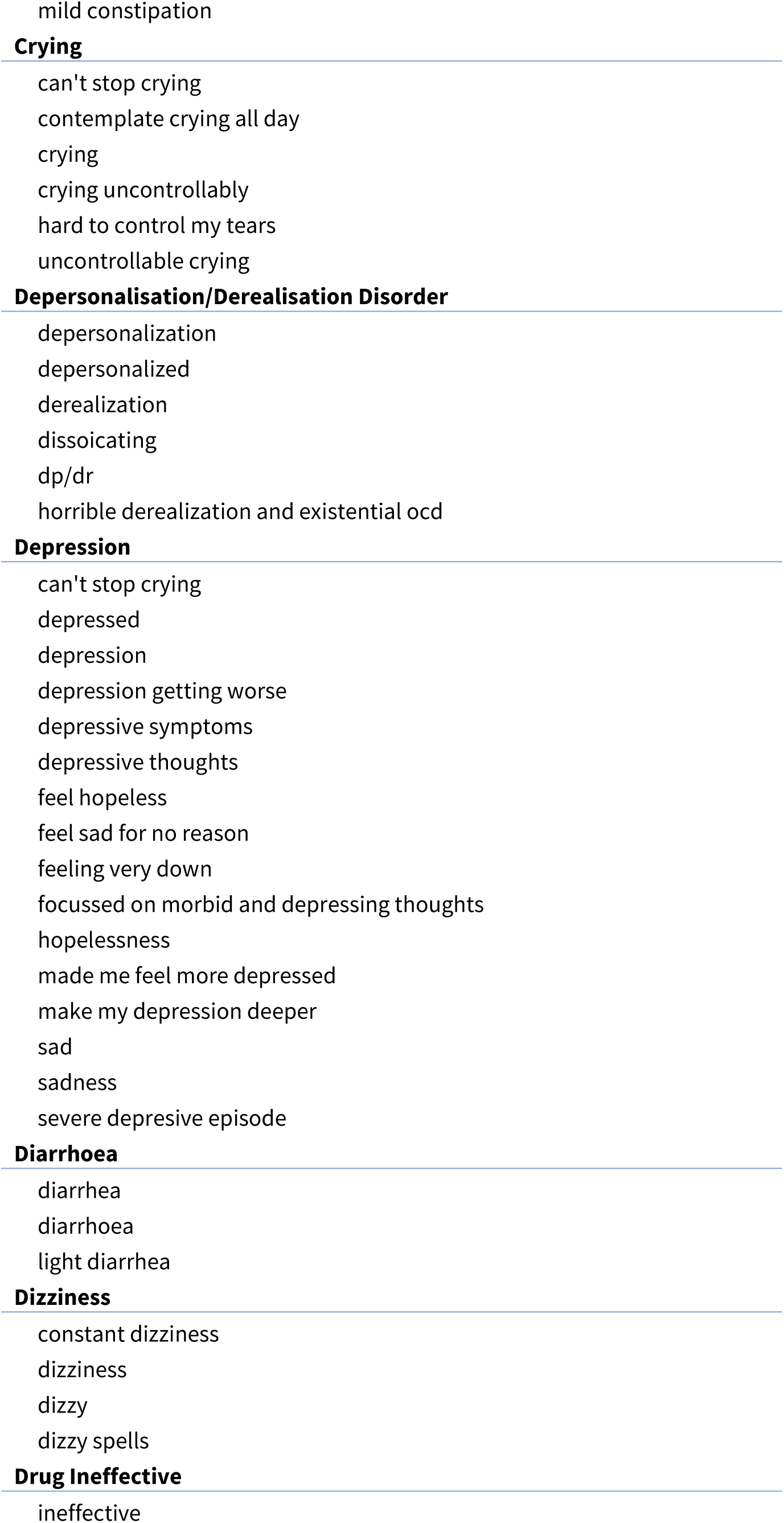
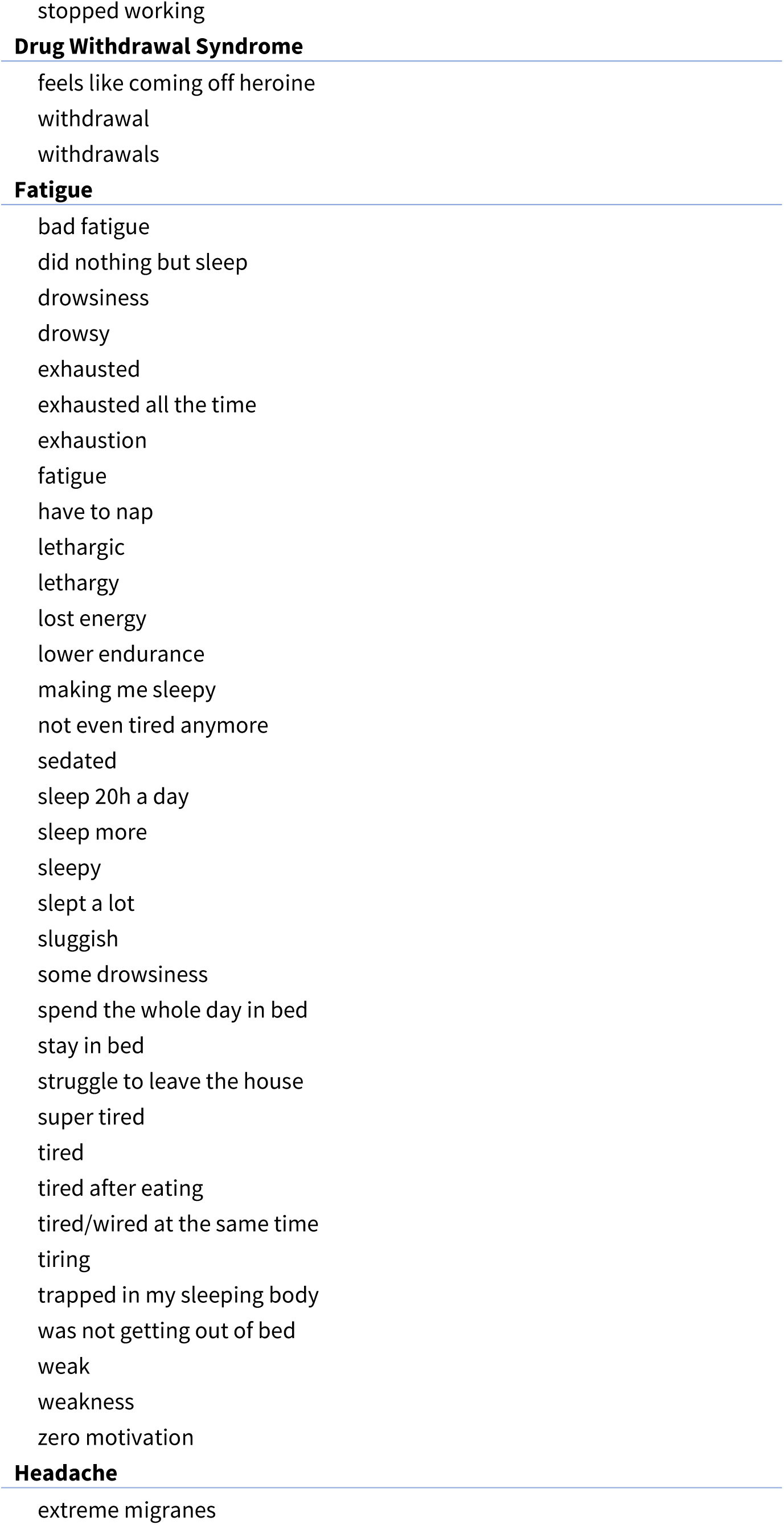
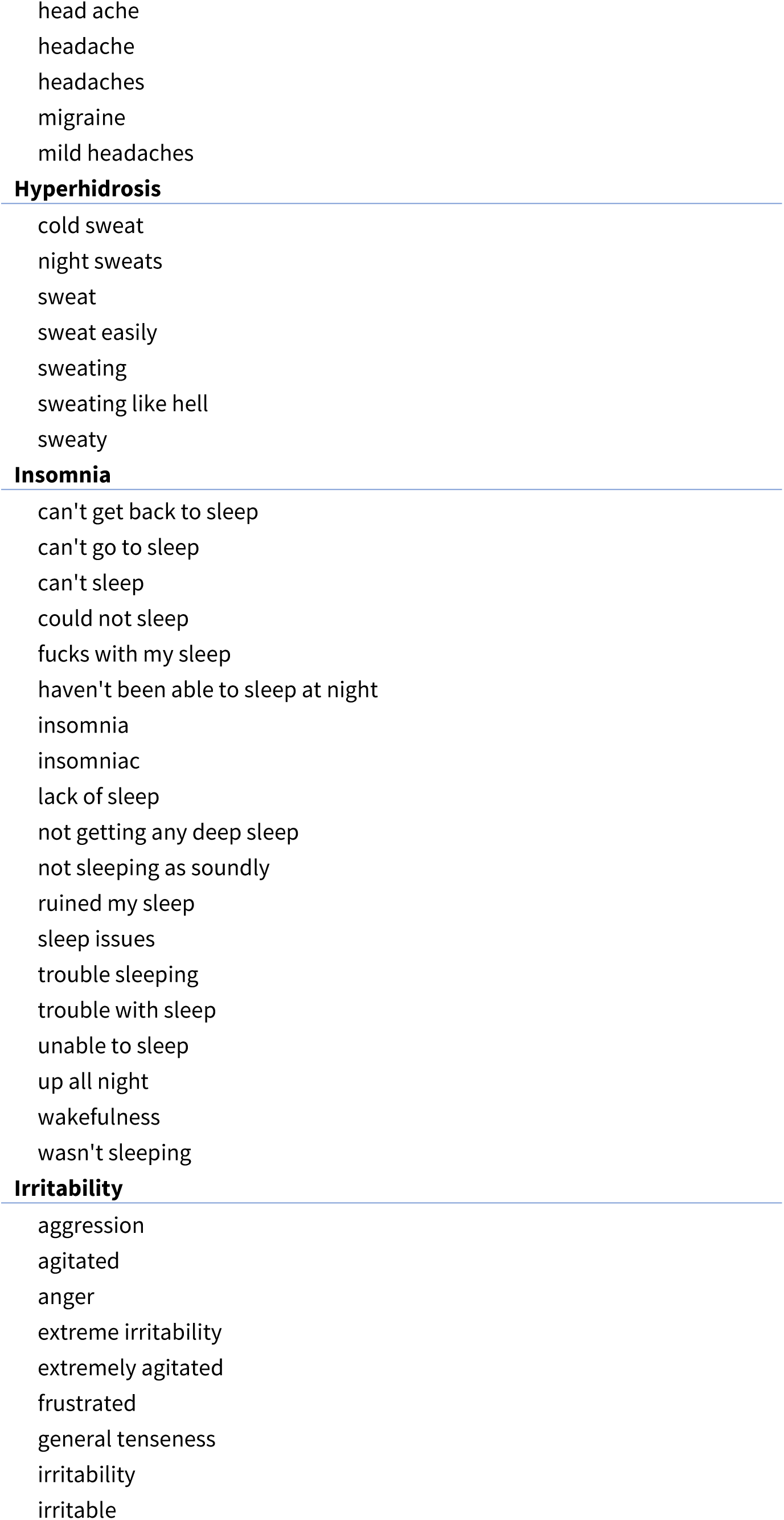
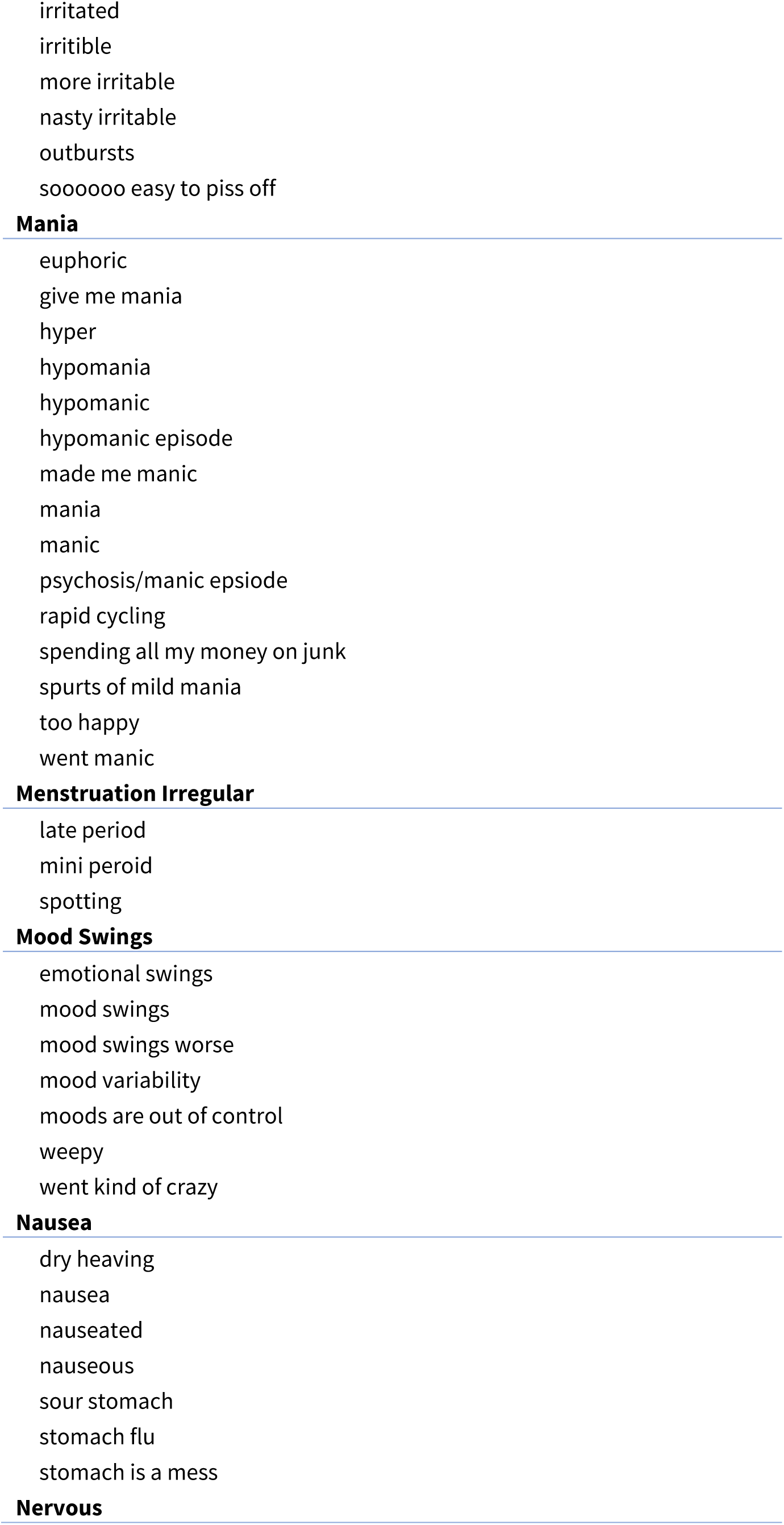
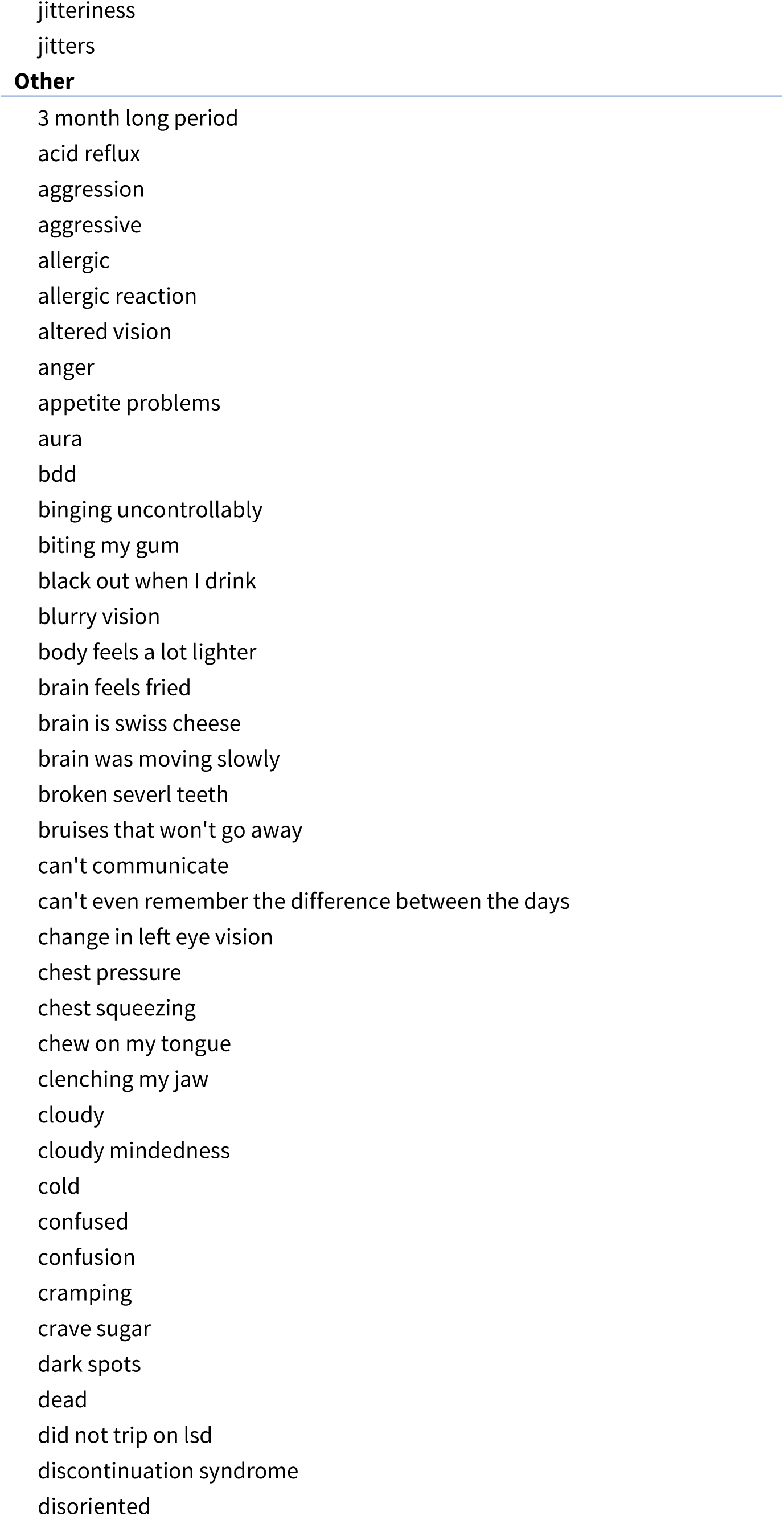
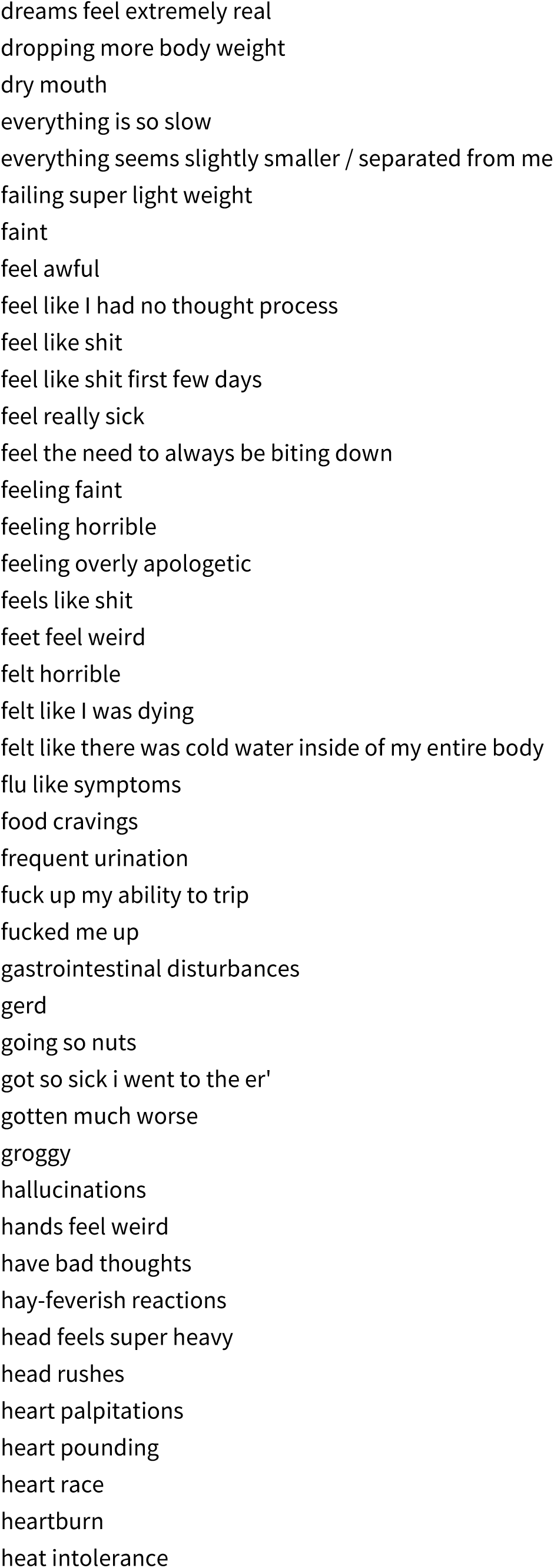
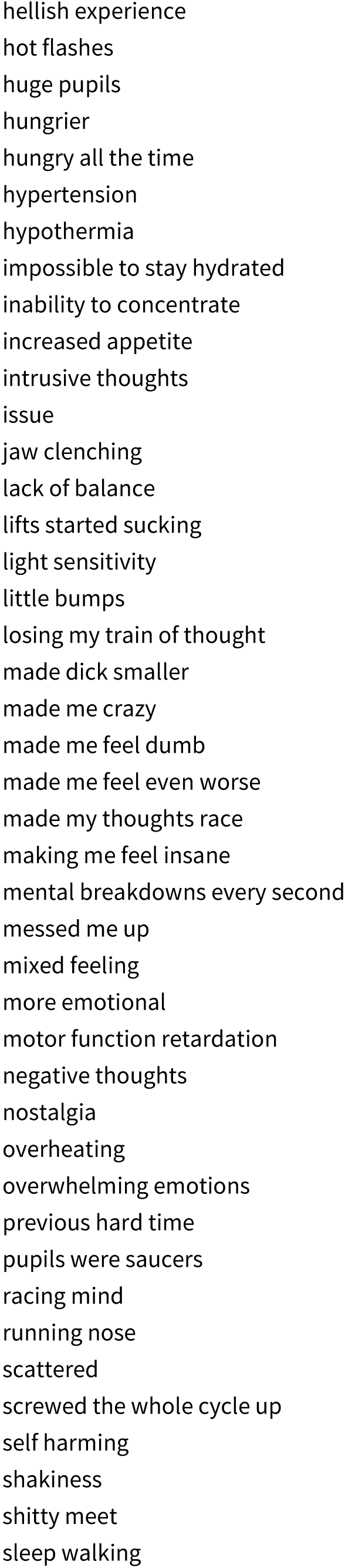
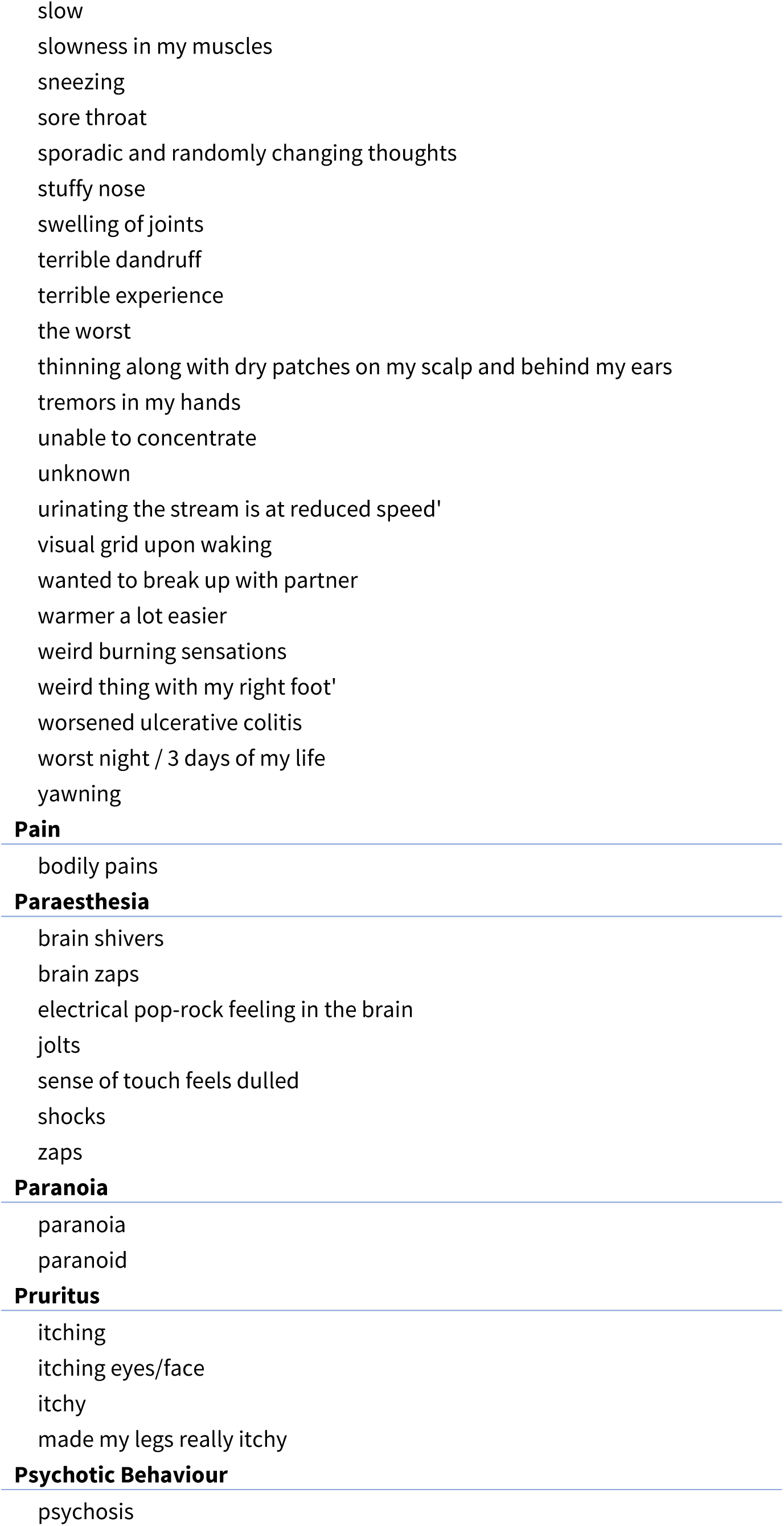
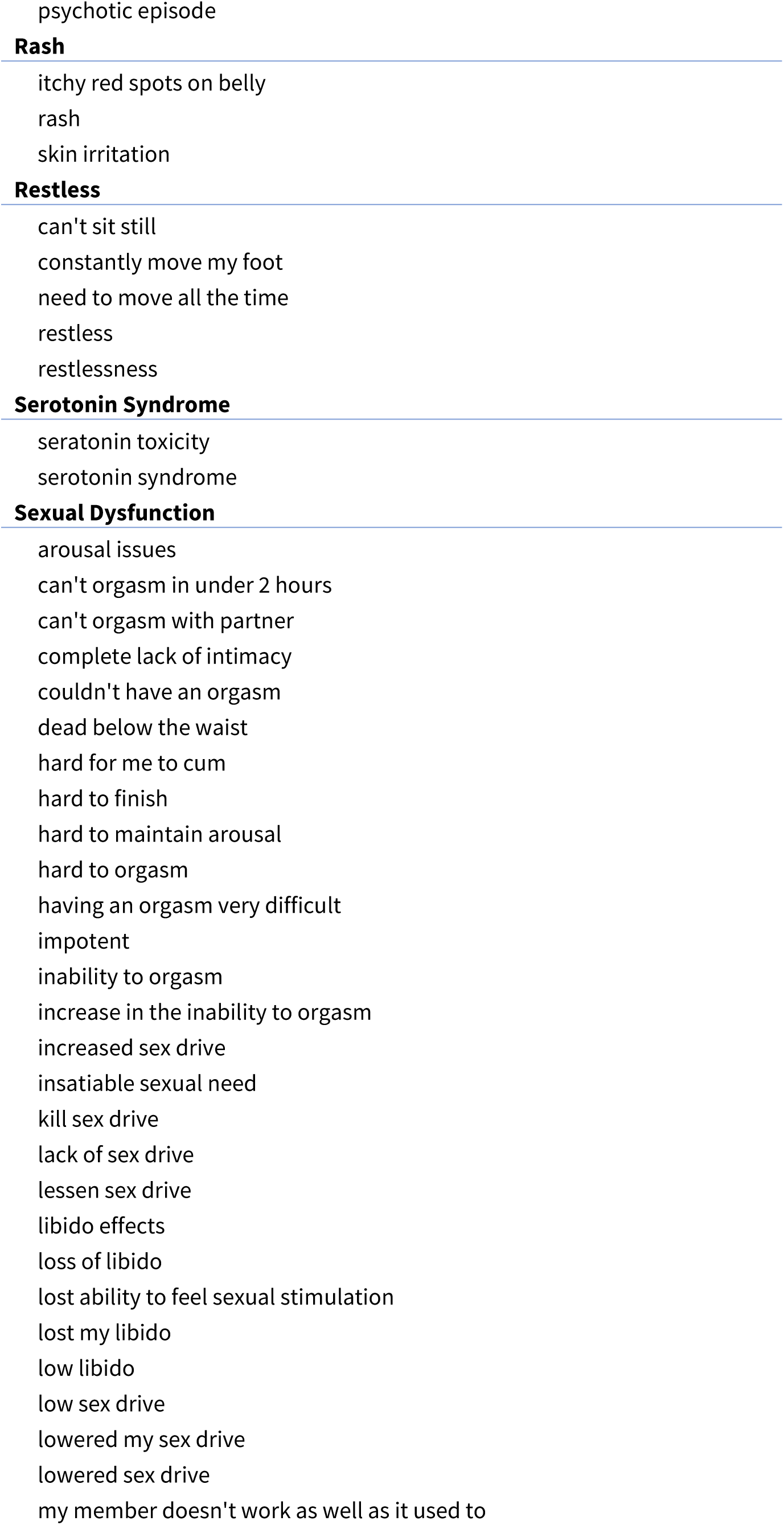
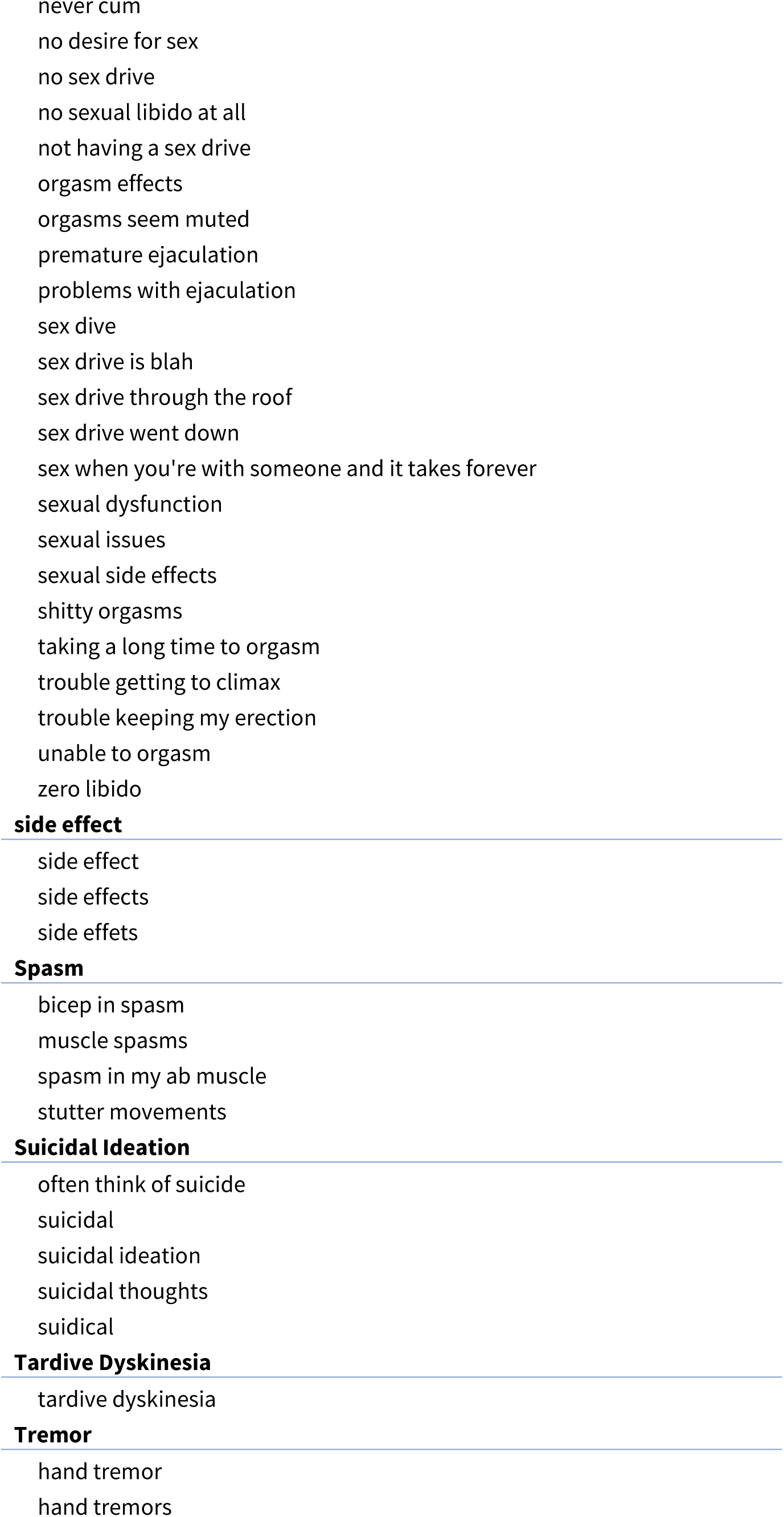
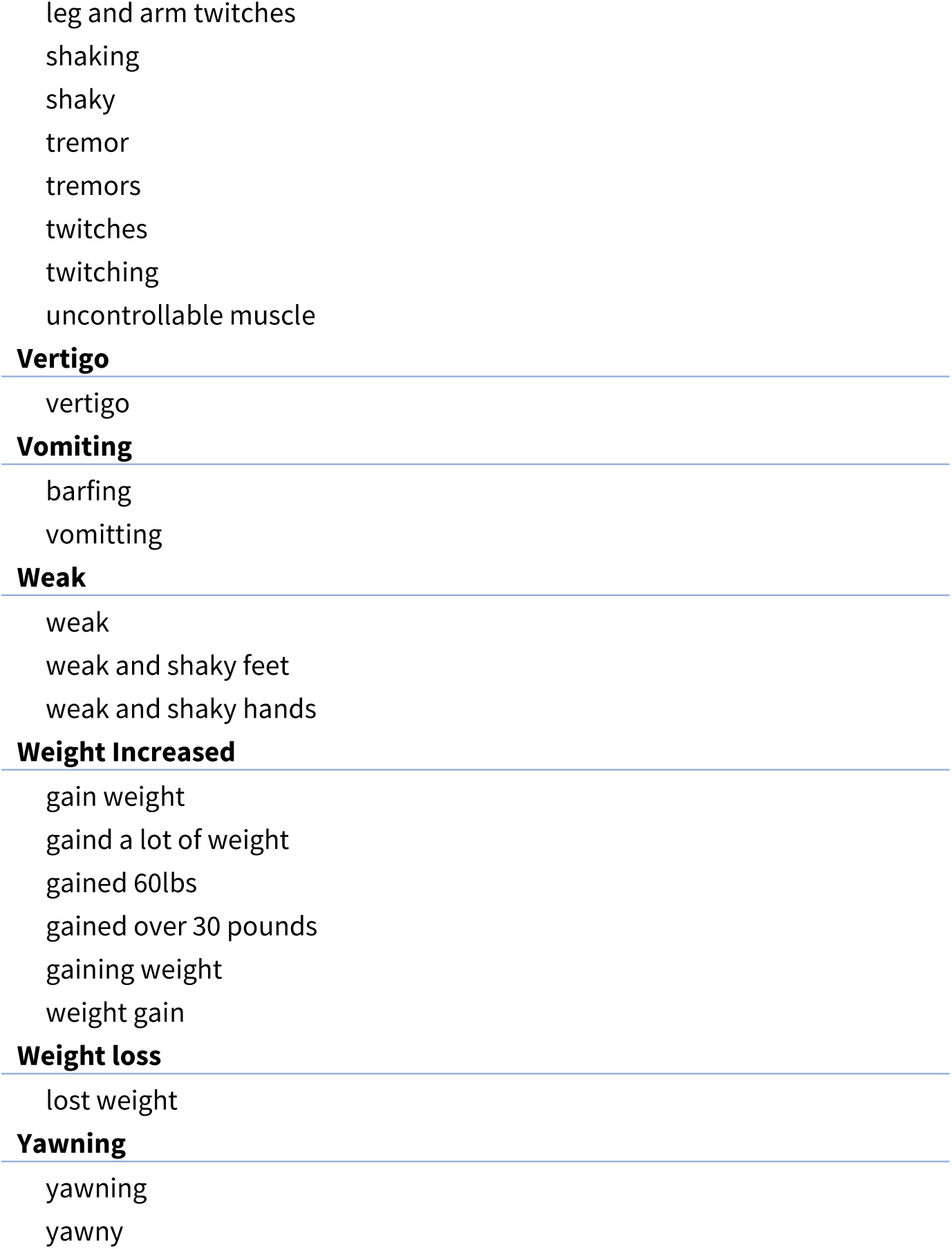

## Appendix 2: Visualization of Reddit and FAERS Adverse Event Rates

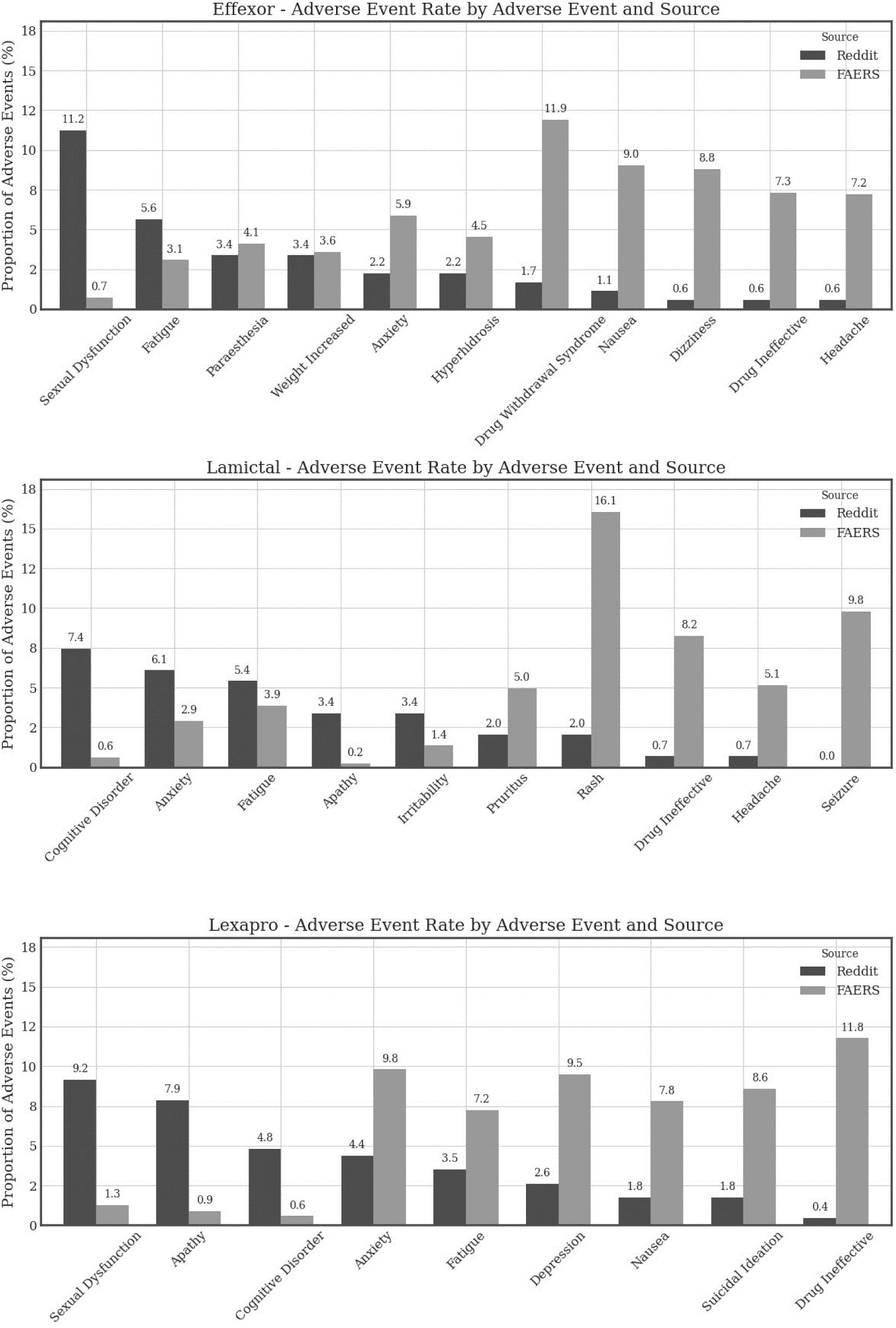
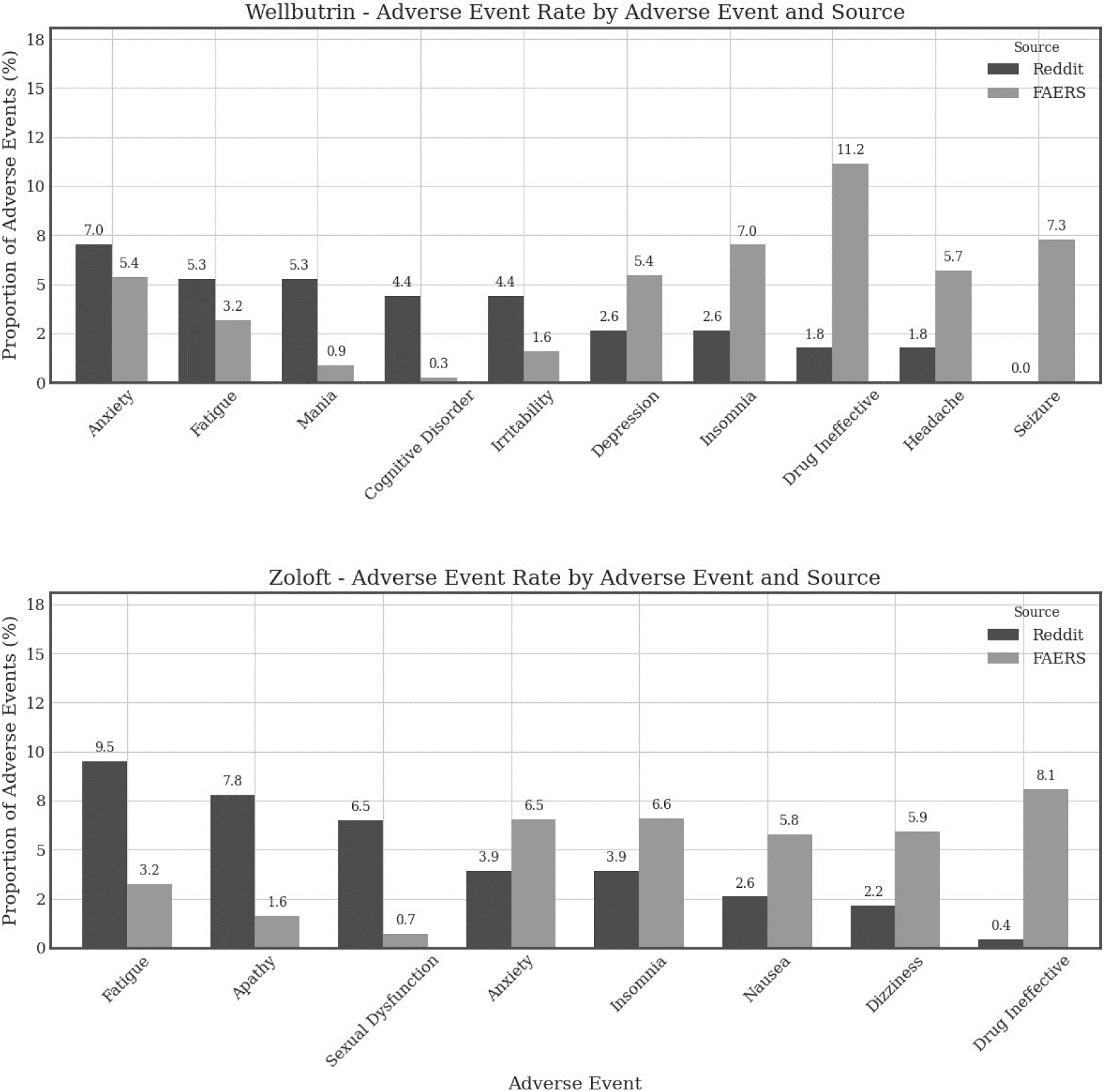

## Footnotes

1 The Twitter platform was renamed X in 2023 but this study uses Twitter’s original name to avoid confusion.

1 https://www.iqvia.com/insights/the-iqvia-institute/available-iqvia-data

## Notes

### Competing Interest Statement

The authors have declared no competing interest.

### Funding Statement

This study did not receive any funding.

